# Risk of Corona virus disease 2019 (COVID-19) among spectacles wearing population of Northern India

**DOI:** 10.1101/2021.02.12.21249710

**Authors:** Amit Kumar Saxena

## Abstract

**Introduction:** Severe Acute Respiratory Syndrome Corona virus-2 (SARS-CoV-2) spread mainly through respiratory droplets and contact routes. Long term use of spectacles may prevent repeated touching and rubbing of the eyes. Aim of the study is to find out the protective effectiveness of the spectacles against COVID-19, if present.

**Objectives:** To know the association between infection with SARSCoV-2 and wearing of spectacles.

**Materials and methods:** In this study, 304 patients of Corona virus disease 2019 (COVID-19) were selected. Their spectacles wearing behaviour were assessed through a questionnaire. Spectacles wearing behaviour of general population were obtained from older studies (for comparison). Data was put in the tabulate form and Chi- Square test was used for statistical analysis.

**Results:** In this study, among the 304 total patients, 58 patients showed the behavior of using spectacles continuously during day time and always on outdoor activities. While the spectacles wearing behaviour is about 40% among general Indian population. The protective effectiveness of the spectacles was found statistically significant (p-value. 00113).

**Conclusion:** The present study showed that the occurrence of Covid-19 was less in spectacles wearing population than the population not wearing those. The nasolacrimal duct may be a route of virus transmission from conjunctival sac to the nasopharynx.

## Introduction

Corona virus disease 2019 (COVID-19) is a major disaster to mankind this year. World Health Organization (WHO) declared it a pandemic on March 11, 2020[1]. It is a respiratory and vascular illness caused by a virus named Severe Acute Respiratory Syndrome Corona virus-2 (SARSCoV-2) of beta-corona virus family. It is a single strand RNA enveloped virus [2]. This virus spread from animal to person and person to person through respiratory droplets and contact route [3]. Coughing, sneezing and even talking by an infected person produce large droplets and small aerosols. Droplet transmission is a major route of infection. Droplets can infect a nearby healthy person while aerosols are dispersed in the air and inhalation of this aerosol can infect the person from a distance [4]. When a healthy person comes in close contact with a sick person, his nasal, oral and conjunctival mucosa are exposed to the virus containing respiratory droplets. Some of the studies show that the virus can survive on the surface of different objects for days [5], [6]. Transmission through contact occurs by touching the face, nose, mouth and eyes after direct contact and fomites used by the infected person [7]. This type of transmission in which person’s own contaminated hands make subsequent contacts with other body parts and introduce that contaminated material to those body parts, is also known as self-inoculation[8]. It is observed that all the respiratory infection virus can transmit by self-inoculation. An individual has habit of touching his own face on average 23 times in an hour[9] and his eyes on average 3 times per hour[10].

Prevention is always better than cure. Government of India issued an advisory asking every person to wear the face mask in April 2020, during the sudden spike in COVID-19 cases [11]. Centers for Disease Control and Prevention (CDC) also advised people to avoid close contact, maintained hand hygiene and use face mask to reduce the spread of the virus [12].

Wearing the face mask properly by a healthy person can reduce the risk of virus infection by checking the entry of the virus containing droplets and aerosols into the nose and mouth. It also reduces the number of respiratory droplets and aerosols coming from the nose and mouth of the infected person during coughing, sneezing or talking. Face mask also protects us from self-inoculation of the virus. Touching one’s nose and mouth is significantly reduced when wearing a face mask properly [13]. But wearing a face mask does not protects the eyes from getting the viral infection through respiratory droplets and self-inoculation. The conjunctival mucosa may be the initial site of infection because it is directly exposed to external pathogens. And angiotensin converting enzyme-2 (ACE-2) receptors are also present on it [14]. SARSCoV-2 virus enters host cells via the ACE-2 receptors [15]. Besides nasolacrimal duct may be another route which can transfer the virus from the conjunctival sac to the nasopharynx [16], [17], [18], [19]. The mucosa of the conjunctival sac is in continuation with that of the upper respiratory tract through the nasolacrimal duct. So it is advised that the health care workers should use face shields and goggles to protect their eyes. Wearing the spectacles do not protect the eyes as much as the goggles, yet it may provide some degree of protection.

The prevalence of refractive errors in India is 53.1 % in adults, and 10.2 % of adults have uncorrected refractive errors (not using spectacles) [20]. From the data described above, we can estimate that about 40 % of Indian persons use spectacles. Aim of this study is to know the association between infection with SARSCoV-2 and wearing of spectacle. It was an attempt to find out the protective effectiveness of the spectacles against COVID-19, if present.

## Materials and methods

During the month of September 2020, India got its peak in COVID-19 cases with about one lakh cases in a day. This study was conducted from August 26, 2020, to September 8, 2020, in KV Covid Hospital, Kanpur Dehat, a district of Northern India. This cross sectional study was approved by the ethical committee KV Covid Hospital, Department of Health, Kanpur Dehat and followed all the guidelines of the declaration of Helsinki. There were total 317 patients admitted in that period. Among them, 304 patients were selected for the study after getting informed consent. The inclusion and exclusion criteria are given below-

### Inclusion criteria

All the admitted patients of mild COVID-19 (SpO_2_ >96%) whose age was more than 10 years, were included in the study. All the patients were Reverse Transcription Polymerase Chain Reaction (RTPCR) positive.

### Exclusion criteria

Children having age less than 10 years were excluded. Patients having moderate and severe disease (SpO_2_ <96%) were excluded from the study. The patients who did not give consent, were excluded from the study.

To know the spectacles wearing behaviour of the patients, a questionnaire was formulated after through discussion among the members of District Blindness Control Society, Kanpur Dehat. The printed version of the questionnaire was not given to the patients due to Covid protocol. It was fulfilled by a single person during history taking and clinical rounds. For testing its reliability, 20 random patients were selected from the sample and fulfilled the questionnaire two times by the same patients, three days apart. Variables were calculated and reliability was measured using ‘test - retest reliability’ method. The test was found reliable (coefficient alpha > 0.7). Validity could not be assessed properly at Covid Hospital using Covid protocols. However, possession and usage of the spectacles by the patients might be a validity factor. (Annexure)

### Annexure

#### Questionnaire

The questionnaire had following points-

1. Do you have proper vision- Yes/ No
2. If not, do you use spectacles- Yes/ No
3. If yes, duration of wearing spectacles-
  a. All the time □
  b. Only while reading □
  c. Occasionally □
4. Do you use sunglasses- Yes/ No
5. If yes, its duration-
  a. Always when go out □
  b. Occasionally □

Data of COVID-19 patients were analysed from the available records for age, gender and spectacles wearing behaviour. Data about spectacles wearing behaviour in general population were not possible during the peak of the epidemic. It was extracted from the earlier studies [20]. From those records, risk of COVID-19 was calculated in long term spectacles wearers as well as in persons not using spectacles. The long-term spectacles wearers were those persons who used spectacles more than 8 hours in a day or using sunglasses always on outdoor activities [21]. The data was put in tabulate form. Statistical analysis was done using Chi-Square test with p value <.05 as significant. The software used was Statistical Package for Social Sciences (SPSS) version 21.0 for windows.

## Results

a. ***Demographic profiles of the patients-*** Data of 304 patients was collected. There were 223 male and 81 female patients. Detailed age and sex wise description of patients was shown in the Table 1. The mean age of the patients was 39.89 ± 16.43 years with range 10-80 years.
b. ***Spectacles wearing behaviour of the patients-*** After analysis of the questionnaire data, following results were obtained-
  1. 236 patients answered that their vision was correct and did not require spectacles. 68 patients admitted that their vision was not proper. In these 68 patient, 60 patients had spectacles and used those.
  2. Amongst the above 60 patients, 42 patients responded that they used the spectacles all the time. They were considered as long term spectacles wearers. 18 patients told that they did not use the spectacles all the time. They used those while reading or doing close work. So they were not considered as long term spectacles wearers.
  3. 193 patients told that they used sunglasses, in which 16 patients used it always on outdoor activities and 177 patients used it occasionally. In this study, those 16 patients were considered as long term spectacles wearers.

**Table 1.** Demographic profile of the patients included in the study.

| Age group (in years) | Male patients | Female patients | Total |
| --- | --- | --- | --- |
| 10- 40 | 112 | 43 | 155 |
| 40- 60 | 73 | 36 | 109 |
| >60 | 28 | 12 | 40 |
| Total | 213 | 91 | 304 |

In this study, total 58 (42+16) patients showed the behaviour of using spectacles continuously during day time and always on outdoor activity. This figure was about 19% of persons taken in the case sample. In other words, 19% population of sample size 304 patients used spectacles most of the day time (>8 hours) and considered as long term spectacles wearers. The spectacles wearing behaviour in general population was described earlier in introduction and methodology section. From the earlier studies, it is estimated that about 40 % of Indian persons use spectacles [20]. In brief, infection with SARSCoV-2 virus and spectacles using behaviour was summarized in table-2.

**Table 2.**
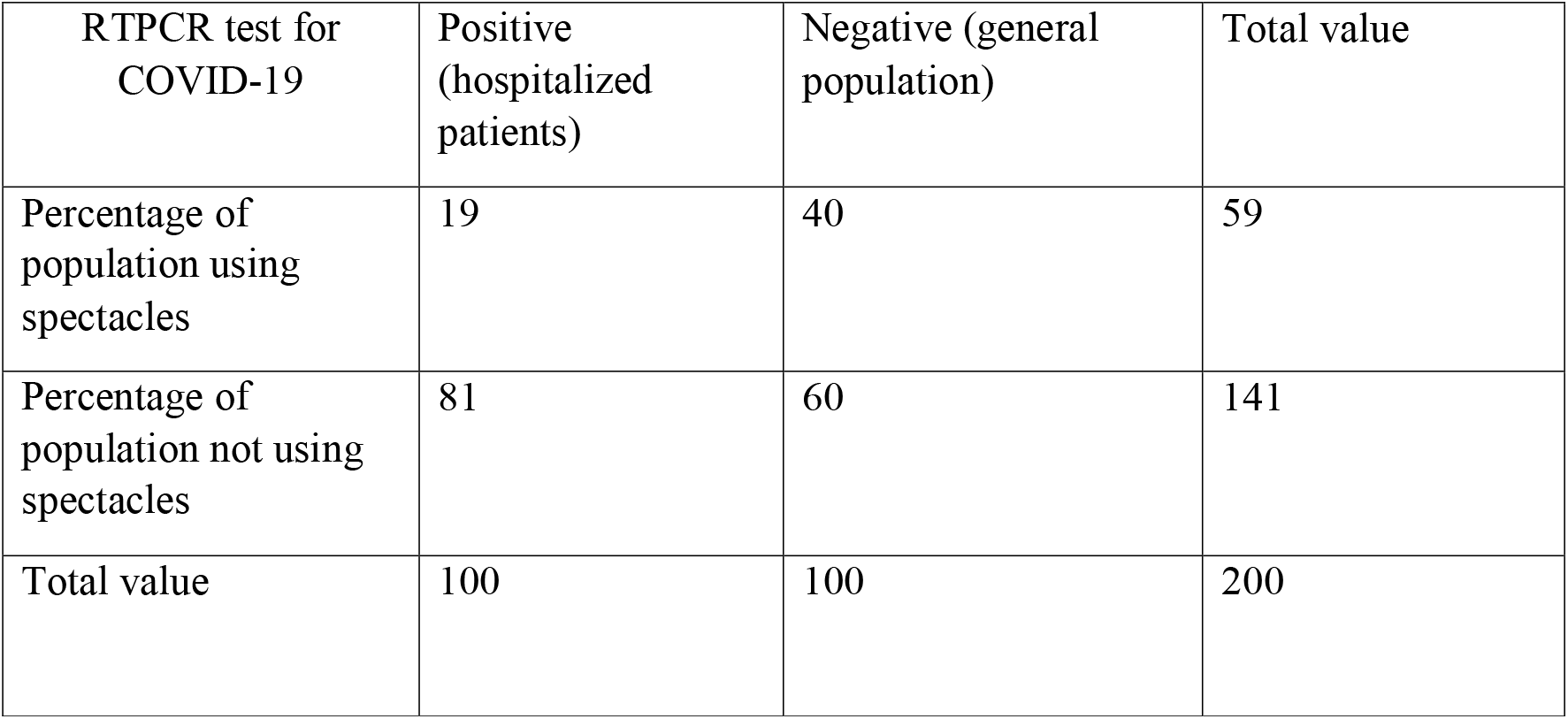
Infection with SARSCoV-2 virus and spectacles using behaviour of persons.

Chi-Square test was used to know the association between infection with SARSCoV-2 virus and spectacles wearing behaviour. The association was found statistically significant having p-value .00113.

## Discussion

To reduce the global burden of COVID-19 cases, we shall have to find out every possible method of virus transmission. We should take all the precautions and measures to reduce its transmission. In this study, risk of the disease in long term spectacles wearers was 0.48 as compared to 1.36 in population not using spectacles. The risk ratio was 0.36 which means that the risk of disease was 2-3 times less in spectacles wearing population. The study was also found statistically significant having p-value .00113. W Zeng et al. showed that long term wearing of eyeglasses (> 8hours/ day) might reduce the susceptibility to COVID-19 disease [21]. It might be due to less touching and rubbing of eyes while wearing spectacles. The study done by Kwok YLA et al. showed that touching our face is a frequent habit and on average every normal person touches his face 23 times and his eyes about 3 times in an hour[10].

Earlier studies showed the presence of SARSCoV-2 virus RNA in conjunctival sac. The RNA was detected in conjunctival swabs of 24% of patients of COVID-19 in a study conducted by Arora R et al. [22]. There are some case reports in which conjunctivitis was the only presentation of COVID-19 [23]. Shengjie Li et al. showed that ACE-2 receptors are present on the human conjunctiva, especially on conjunctival epithelial cells. They found that COVID-19 can be transmitted through the conjunctiva [24]. However Lange C et al. gave the opposite concept. They showed that conjunctival ACE-2 receptors have very limited role in transmission of the disease [25]. The study done by JJ Miner et al. in November 2020 showed that SARSCoV-2 virus do not replicate in human corneal explants [26]. They showed that infection of corneal tissue was extremely rare. However nasolacrimal duct is another route which opens the passage of the virus from conjunctival sac to nasopharynx. Some studies asserted that the virus may pass from conjunctiva to nasopharynx through the nasolacrimal duct [16], [17], [18], [19]. So it is advised that the health care workers should use eye safety goggles when coming into contact with patients of COVID-19.

## Limitations

There are some confounding factors in the present study as follow of preventive measures, education status of the patients etc. The uneducated and lower socioeconomic persons are more prone to get the infection. They do not follow the preventive guidelines properly. They also have habit of using spectacles less than the educated persons. The sample size was small. The period of the study was also short. The spectacles wearing behaviour of general population could not be measured during the peak of epidemic. More studies should be done to know the effect of using spectacles on the epidemiology of COVID-19.

## Conclusion

This present study showed that the occurrence of COVID-19 was less in spectacles wearing population than the population not using spectacles. Protective role of the spectacles was found statistically significant, if those were used for long period of the day (>8 hours). Touching and rubbing of the eyes with contaminated hands may be a significant route of infection for SARSCoV-2 virus.

## Data Availability

Data available at record section KV COVID Hospital, Department of Health, Kanpur Dehat.

## Acknowledgement

I am very thankful to Dr. A P Verma, Nodal Officer, KV Covid Hospital Kanpur Dehat for helping me in this study. I am also thankful to the members of District Blindness Control Society, Kanpur Dehat for formulation of the questionnaire.

